# The Madrid Manic Group (MadManic) Cohort: Multi-Omics and Digital Phenotyping For the Studies of Severe Mental Disorders and Suicidality

**DOI:** 10.64898/2026.04.14.26350865

**Authors:** Inés García-Ortiz, Raúl Somavilla Cabrero, Elisabet Madridejos Palomares, Miriam Martínez-Jiménez, Rosa Ana Bello Sousa, Inés Carpio-López, Sergio Sánchez-Alonso, Sergio Benavente López, Laura Mata-Iturralde, Raquel Álvarez García, Diego Romero-Miguel, Laura Jiménez Muñoz, Ezequiel Di Stasio, Ana José Ortega Heras, Silvia de la Fuente Rodríguez, Irene Aguilar Castillo, Ana Lara Fernández, Irving Clarke Gil, Concepción Vaquero Lorenzo, Per Hoffmann, Carlos López de la Hoz, Noelia Borge García, José Abad Valle, María Juncal Sánchez Alonso, Elena Arroyo Bello, Rebeca Jiménez Peral, Ana María de Granda Beltrán, Janice M. Fullerton, Marta Bermejo Bermejo, Lucía Albarracín-García, Enrique Baca-García, Claudio Toma

**Author notes:** Corresponding author: Claudio Toma, Centro de Biología Molecular (CBMSO), Calle Nicolás Cabrera 1, 28049 Madrid, Spain. These authors contributed equally (Enrique Baca-García, Claudio Toma).

## Abstract

Severe mental disorders (SMDs), including bipolar disorder, schizophrenia, and major depressive disorder, are highly complex conditions associated with a substantial clinical burden and an increased suicide risk. Here, we present the Madrid Manic Cohort (*MadManic*), a large-scale initiative from Spain designed to integrate genomic, multi-omics, clinical, and digital phenotyping data to investigate the biological basis and clinical heterogeneity of SMDs. The cohort is still expanding and currently includes over 4,400 participants (∼2,300 psychiatric patients and ∼2,100 controls) and >11,000 biospecimens. Genotyping, transcriptomic and epigenetic data are available for different subsets of the cohort.

By establishing the *MadManic* cohort we aim to integrate molecular data with detailed clinical and longitudinal digital information, allowing a more precise characterization of patient subgroups based on biological and phenotypic profiles. The *MadManic* cohort is well positioned to contribute to major international efforts in psychiatric genetics by enhancing the representation of Southern European populations, and advancing the identification of genetic risk, clinical predictors, and pharmacogenomic markers of treatment response.

This cohort represents a valuable resource for advancing precision psychiatry, with the potential to improve risk prediction and guide personalized interventions in SMDs.

## INTRODUCTION

Severe mental disorders (SMDs) encompass a spectrum of chronic and debilitating psychiatric conditions, including bipolar disorder (BD), schizophrenia (SCZ), and persistent major depressive disorder (MDD). These mental conditions are associated with significant functional impairment, frequent psychiatric hospitalizations, and elevated suicide behaviour (SUI)^1,2^. SMDs are often comorbid with other psychiatric traits or conditions, including substance use disorder (SUD), and personality disorders, amongst others^3,4^. BD and SCZ are chronic and severe mental illnesses affecting approximately 1% of the global population^5,6^. BD is characterized by recurrent episodes of mania, depression, and mixed mood states, and is often accompanied by psychosis^5^. Around 50% of individuals diagnosed with BD, attempt suicide at least once in their lifetime, with an estimated 15-17% ultimately dying by suicide^7^. SCZ is associated with severe cognitive, emotional, and functional disturbances^8^. Suicide rates among individuals with SCZ are significantly elevated, with an estimated of ∼10% dying by suicide^9^. MDD is a common and heterogeneous psychiatric condition with a lifetime estimated prevalence of ∼10-15%, characterized by persistent low mood, loss of interest, and impaired daily functioning^10^. SMDs carry a substantial burden on individuals, families, and healthcare systems, necessitating the development of novel approaches for diagnosis, treatment, and prevention of adverse outcomes. Suicide is one of the most devastating consequences of SMDs, with approximately 90% of suicide deaths occurring amongst individuals with psychiatric disorders^11^. In Spain, suicide is the leading cause of unnatural death, doubling those for traffic accidents, with rates remaining stable over the past two decades (∼3,500-4,000 deaths annually)^12^. The high prevalence of suicide amongst individuals with BD, SCZ, and MDD highlights the critical need for targeted interventions and personalized risk assessment strategies.

Despite advances in pharmacological and psychotherapeutic treatments, SMDs remain major contributors to psychiatric hospitalizations and healthcare expenditures. Traditional clinical instruments that rely on face-to-face assessments exhibit limited predictive power in preventing suicide and reducing hospital admissions. Mobile health (mHealth) technologies and ecological momentary assessment (EMA) offer promising alternatives by facilitating real-time monitoring of mood states, behaviours, symptoms, and suicide risk in patients with SMDs^13^. Through smartphone applications, EMA enables the collection of longitudinal data with high precision, allowing for personalized interventions and improved symptom tracking^14^. However, while digital phenotyping holds great potential for suicide prevention, its clinical implementation remains limited.

Beyond real-time monitoring, genetic susceptibility plays a significant role in the pathophysiology of SMDs and suicidality^15^.

Psychiatric phenotypes, including SMDs and suicidality are highly polygenic conditions, that result from a complex interplay between hundreds of risk alleles from common variants or single nucleotide polymorphisms (SNPs) and several pathogenic rare variants or single nucleotide variants (SNV)^16,17^. Additionally, other classes of genetic variation, such as *de novo* variants and copy number variants, usually confer larger effect sizes to the disease risk^17,18^. Genome-wide association studies (GWAS) meta-analyses performed by the Psychiatric Genomics Consortium (PGC) have identified multiple common variants associated with several psychiatric phenotypes. Furthermore, next-generation sequencing (NGS) technologies, such as whole exome sequencing (WES) and whole genome sequencing (WGS), have begun to uncover rare genetic variants that contribute to susceptibility of SMDs^19^. The largest GWAS of BD to date, conducted by the PGC, identified 298 significant loci using data from over 158,000 cases and 2.8 million controls^20^. Similarly, large-scale genomic studies from the PGC have identified hundreds of loci associated with SCZ and MDD ^21,22^. Emerging genomic research into suicidality further supports the notion of specific genetic contributions to suicide ideation (SI), suicide attempt (SA), and suicide death (SD)^23,24^. Despite these global efforts have enhanced gene discovery, larger cohorts are needed to increase statistical power for gene and variant detection. However, GWAS are not only useful to target biological mechanisms implicated in these complex disorders, but provide also a valuable tool for estimating individual genetic liability through the Polygenic Risk Scores (PRS), offering a potential avenue for integrating genomic data into personalized psychiatric care or risk estimation^25^.

Genetic variations significantly influence not only disease risk, but also individual response to psychiatric medications, including antipsychotics, antidepressants, and mood stabilizers. Pharmacogenomic studies have identified genetic markers that affect drug metabolism, efficacy, and potential adverse effects. Variants in the *CYP2D6* and *CYP2C19* genes impact the metabolism of antidepressants and antipsychotics, altering plasma drug levels and treatment outcomes^26^. Lithium responders are more frequently observed in families with a high density of BD patients^27^, for which linkage studies have identified several non-overlapping loci involved in lithium response^28^. These family-based studies suggested that long-term lithium response is a heritable trait, which has prompted to several genetic studies. The largest GWAS of lithium response to date was performed by the Consortium on Lithium Genetics (ConLi^+^Gen) in >2,500 BD patients, implicating a single locus on chromosome 21 containing two non-coding RNAs (*AL157359.3* and *AL157359.4*)^29^. Despite this study represents the first large attempt to uncover the genetics for lithium response, it remains largely underpowered due to its limited sample size, and its results cannot be translated to clinical practise yet.

Efforts to translate genetic findings into clinical practice in psychiatry have largely relied on genetic data alone, often disregarding or minimizing complementary approaches such as RNA sequencing (RNAseq), epigenetics, and metabolomics. While GWAS have identified numerous risk variants for psychiatric disorders, their direct clinical utility remains limited.

Integrating multi-omic approaches, including transcriptomic, epigenetic, and metabologenomic profiling, may provide a more powerful framework for understanding disease mechanisms, treatment response, and individual risk. Combining genome-wide assays with clinical digital profiles and longitudinal data will enhance predictive modelling, facilitating the development of personalized interventions in psychiatry.

We introduce the Madrid Manic Cohort (*MadManic*), a novel cohort study designed to integrate digital phenotyping, genomic data, multi-omics layers, and personalized medicine approaches to improve our understanding and management of SMDs and suicidality. This study outlines the cohort design, scientific objectives, and data collection strategies, providing a base for future research studies and collaborations.

## METHODS

### Aims

The *MadManic* group aims to develop a robust framework for personalized medicine in patients with SMDs by integrating EMA through smartphone-based monitoring systems with genomic, transcriptomic, and epigenomic data. Our approach seeks to improve predictive modelling of risk monitoring, diagnostic trajectories, and treatment response, enhancing clinical outcomes and reducing suicide risk. The objectives of future studies will focus on: i) Identify genomic and epigenomic signatures linked to psychiatric disorder risk; ii) Identify pharmacogenetic and pharmacotranscriptomic patterns of treatment response; iii) Develop pioneering and multidisciplinary methods for the identification of psychiatric patients at-high-risk for suicide.

### Study design

The *MadManic* is a novel cohort from Spain initiated in 2022. This cohort, which recruitment is still ongoing, includes patients with BD, SCZ, MDD, SI, SA, SUD, and borderline personality disorder (BPD), and control subjects.

Psychiatric patients are clinically assessed at following hospitals and clinical units in the metropolitan area of Madrid: Hospital Fundación Jiménez Díaz and the related clinical units of Pontones and Quintana, Hospital General de Villalba, Hospital Rey Juan Carlos, and Hospital Infanta Elena de Valdemoro (**Fig. 1**). Controls are recruited at two occupational medicine services of the Consejo Superior de Investigaciones Científicas (CSIC) and at the School of Nursing at Hospital Fundación Jiménez Díaz. Hospitals and clinical units are all based in Madrid (**Fig. 1**). All individuals are required to provide a blood sample for extractions of DNA, RNA, plasma, and serum, or alternatively provide a saliva sample for DNA extraction. Fresh blood samples are processed in 2-6h into biospecimens at Centro de Biología Molecular Severo Ochoa (CBMSO) (**Fig. 2**). After the informed consent, patients are scheduled for an interview with clinical psychologists, who follow a digital clinical protocol and invite the patients to install the mobile apps for the digital phenotyping. Clinical data both from the clinical protocol and the apps are then managed and digitally stored (www.memind.net).

**Figure 1.**
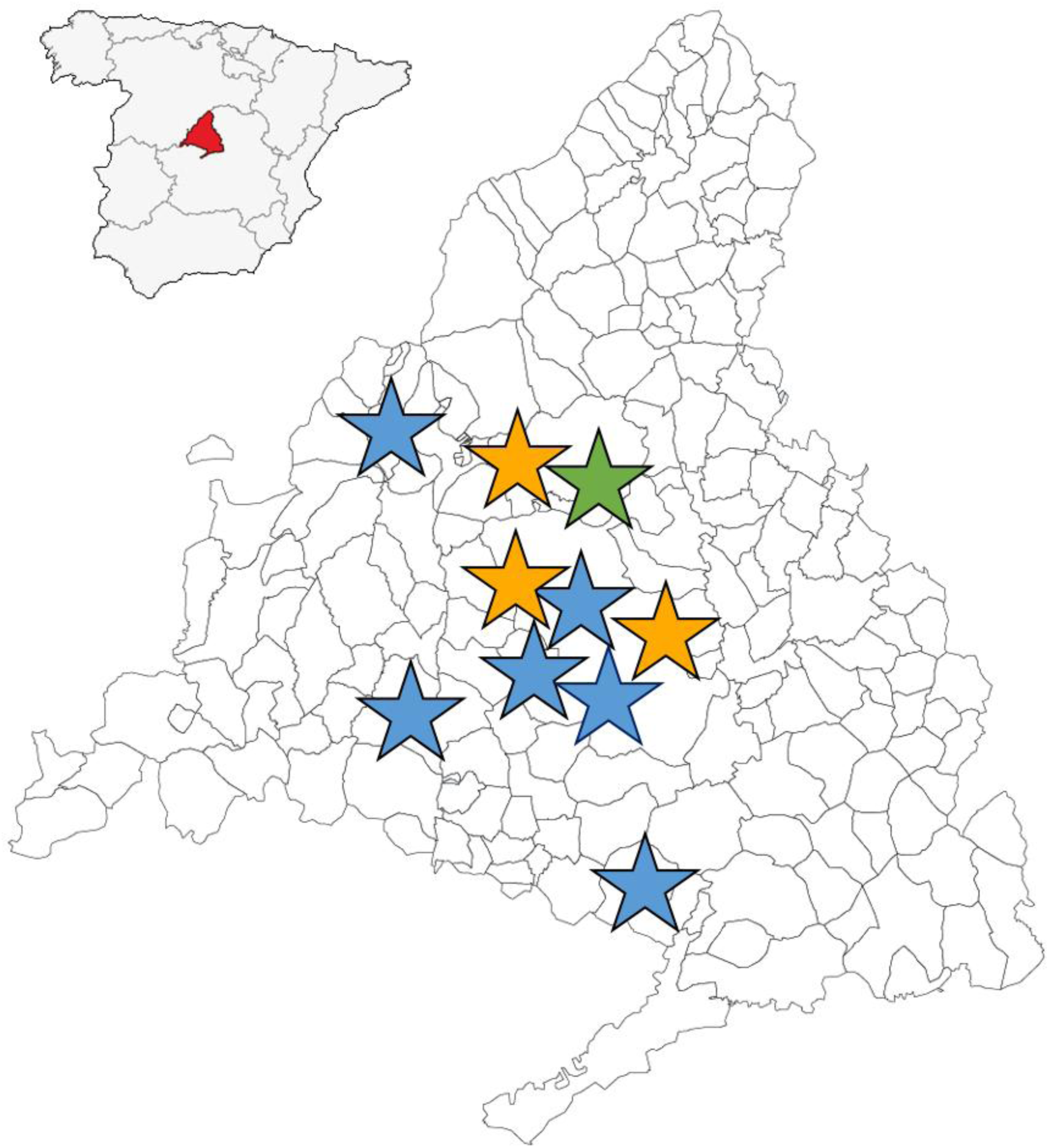
Location of recruitment units in *MadManic* group. The network includes nine sites. Psychiatric patients are assessed in four psychiatric hospitals and two psychiatric units (blue stars) covering a million of Spanish residents. Controls are sourced from donors at occupational medicine services of CSIC and the Nursery School at University Hospital at Fundación Jiménez Díaz (orange stars). All blood samples are processed and stored at CBMSO (green star).

**Figure 2.**
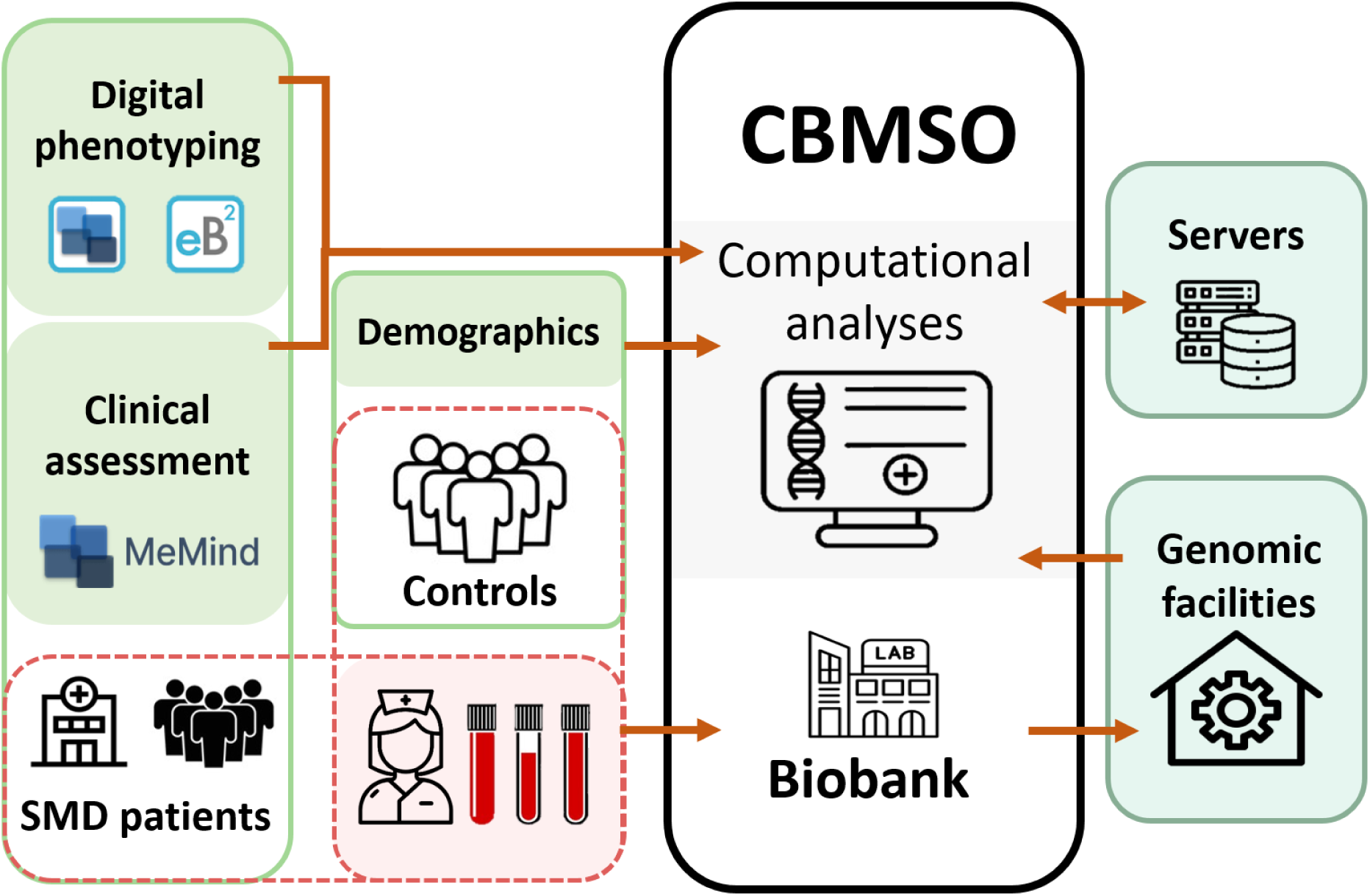
Complete workflow of the *MadManic* group. All the aspects related with clinical records are highlighted in green. The clinical data are managed and stored by the MeMind server. Blood extractions in dashed lines are carried out both for SMD patients and controls, and processed freshly at CBMSO. High-throughput arrays are outsourced at genomic facilities and raw data are sent back to the CBMSO for downstream analyses using computational servers associated to the CBMSO.

### Ethics

Written informed consent is obtained from all the participants prior to their inclusion in the study. Approval to handle clinical, digital, and genomic data of patients and controls and for all aspects of the project was obtained by the research ethics committee of the CSIC (13/2021; 109/2023; 030/2025) and Fundación Jimenez Díaz Hospital (03/06; 15/21; 11/23).

### Clinical assessment and digital phenotyping

Clinical assessments are based on face-to-face interviews collecting clinical data and scales recorded electronically and a digital phenotyping through mobile apps (**Fig. 2**). The clinical protocol has been designed for deep phenotyping of patients with BD and controls (**Table 1**). The protocol includes information regarding three parts. The first part of the protocol includes the following items: i) socio-demographics: ethnic background, work status, level of studies; ii) biometric data: body mass index, waist-to-height ratio, and physical activity to assess obesity related risk factors; iii) sleep habits: insomnia and hypersomnia; iv) substance use: tobacco, coffee and alcohol consumption, consume and abuse of cannabis, cocaine, and other drugs; v) disease trajectory: diagnosis following the ICD-10 codes, bipolar disorder spectrum subphenotype (BD type-I; BD type-II; schizoaffective disorder; other type of bipolar disorder), start of the affection status, start of the first treatment, polarity and modality of the first episode, principal polarity; total number of manic and hypomanic episodes, age at first manic or hypomanic episodes, age of the latest manic or hypomanic episodes, features of the manic or hypomanic episodes; total number of depressive episodes, age at first depressive episodes, age of the latest depressive episodes, features of the depressive episodes; total number of mixed episodes, age at first mixed episodes, age of the latest mixed episodes, features of the mixed episodes; history of psychosis and substance-induced psychosis, and relationship with the mood state; euthymic state or current episodes at blood extraction; rapid cycling; vi) psychiatric comorbidity and age at diagnosis; vii) total number of hospitalizations: those associated with mania, depression, psychosis, and those with suicide behaviour; viii) suicidality: history of SA and total number of SA, age at first SA, number of SA in the last year, medication during the last SA, SI and self-injury questions; ix) family history of psychiatric conditions and diagnosis of family members; and x) traumatic events.

**Table 1.**
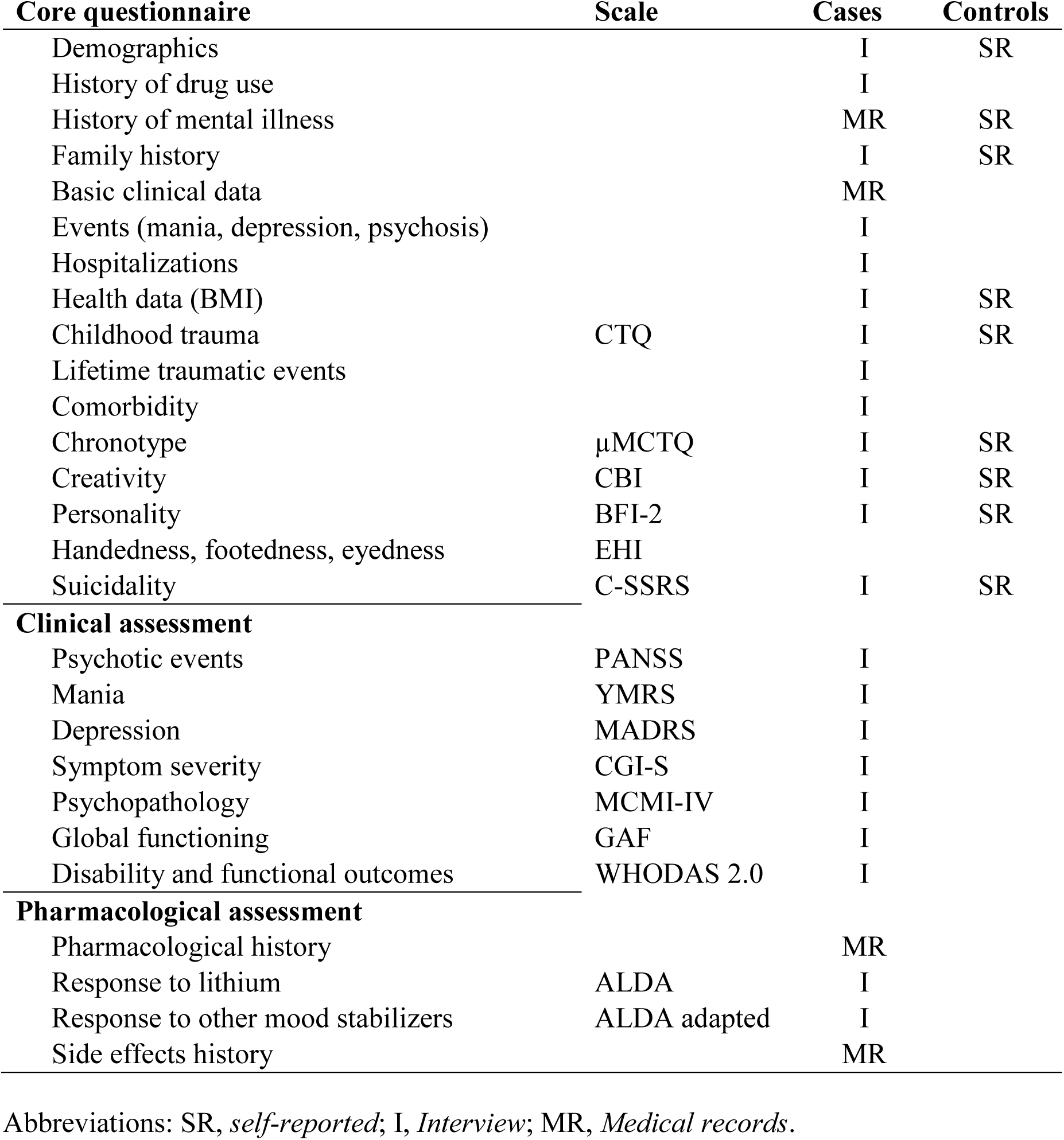
Clinical records and phenotyping scales from participants of the MadManic cohort.

**Table 2.** Collected bioespecimens in the MadManic cohort from psychiatric patients (N: 2,310) and controls (N: 2,111). Psychiatric patients can meet multiple diagnostic categories described below.

|  | **DNA** | **RNA** | **Plasma** | **Serum** |
| --- | --- | --- | --- | --- |
| **Psychiatric subjects** | 2,310 | 1,476 | 1,976 | 230 |
| Bipolar Disorder | 732 | 556 | 556 | 50 |
| Schizophrenia | 356 | 196 | 196 | 100 |
| Suicide Attempters | 1,127 | 627 | 1,127 |  |
| Major Depressive Disorder | 798 | 798 | 798 | 80 |
| Substance Use Disorder | 150 | 150 | 150 |  |
| Borderline Personality Disorder | 142 | 142 | 142 |  |
| **Controls** | 2,111 | 1,431 | 1,431 | 220 |
| **Total** | 4,421 | 2,907 | 3,407 | 450 |

The second part includes pharmacological treatments, including posology, regime, side effects, and response. A complete list of pharmaceutical drugs used from our study group is listed in **Table S1**. The response to lithium is assessed retrospectively using the ALDA scale^27^, a 10-point measure used by Consortium of Lithium Genomics (ConLi^+^Gen) to quantify improvements during long-term treatments with lithium. We have expanded pharmacological assessments adapting the ALDA scale to measure the response to other mood stabilizers, including valproate, carbamazepine, lamotrigine, and oxcarbazepine.

Finally, the third part includes clinical and behavioural scales described in **Table 1**.

The suicide patients are enrolled in the SmartOmics and SMARTcrisis 3.0 projects, assessing suicide-related traits^30–32^. The Columbia–Suicide Severity Rating Scale (C-SSRS) is administrated to assess the severity and immediacy of suicidal ideation and behaviour^33^.

Control subjects complete a structured questionnaire (**Table 1**), including the following two screening questions: (1) “Have you ever received a diagnosis from a psychiatrist of any psychiatric condition?” and (2) “Have you ever been prescribed medication from a psychiatrist to treat psychiatric symptoms such as anxiety, depression, panic attack, etc.?” Only individuals who responded “no” to both questions were included in the control group. SA and SI in the control group are assessed using suitable items identified by the Psychiatric Genomics Consortium Suicide Working Group^23^: i) Diagnostic Interview for Genetic Studies (DIGS) Version 2.0 (Section O, Question 1) to evaluate SA^34^; and MADRS (Section 10 Suicidal thoughts) to assess SI^35^.

### Digital phenotyping

The digital phenotyping is carried out via two smartphone apps, which continuously monitor and register patient’s symptoms through the following systems:

1. *MeMind* (www.memind.net) has been designed to monitor suicide risk and assess suicidality, quality of sleep, negative feelings, etc^13,36^. This application is based on an “active” interaction with patients through random questions along longitudinal periods (weeks/months).
2. Evidence-based Behaviour (eB2) (https://eb2.tech) is based on a passive interaction with patients continuously collecting data via native sensors that register smartphone usage patterns (time of use of the device, phone call duration), mobility (location, distance travelled, and speed), physical activity, sleep quality, and prosody (elements of speech such as intonation, tone, stress, and rhythm)^36,37^.

### Collection of biospecimens

Participants provide a 10 ml peripheral blood sample, which is transported within 1–4 hours from the extraction site to CBMSO for biospecimen processing (**Fig. 2**). Upon arrival, 2.5 ml of the blood sample is used for RNA extraction using the Maxwell® RSC simplyRNA Purification Kit (Promega, Madison, WI). The remaining volume is centrifuged to separate plasma and the buffy coat, the latter used for DNA extraction using the Gentra Puregene Blood Kit (Qiagen, Valencia, CA), following the manufacturer’s protocol.

When blood extraction is not feasible, DNA is alternatively extracted from saliva samples collected using the Oragene ON-600 DNA Extraction kit (DNA Genotek, Ontario, Canada). The DNA concentration of all samples is measured using a NanoDrop spectrophotometer (NanoDrop Technologies, LLC, Wilmington, DE, USA). For the most recent collections, an additional blood sample is used for serum extraction.

The study includes an additional DNA-only collection obtained in a previous recruitment wave (1996–2009), comprising individuals with a history of suicide attempts who had been diagnosed with a psychiatric disorder, as well as healthy controls.

### Genomic arrays

Biospecimens collected from the *MadManic* cohort are currently being used for high-throughput genomic analyses encompassing genotyping, transcriptomics, and epigenetic profiling:

1. Genotyping is performed using the Axiom Spain Biobank Array (Thermo Fisher Scientific), which includes 814,923 markers, being 57,087 quality control (QC) probes. Genotyping of the first wave of this cohort was carried out at the Santiago de Compostela Node of the Centro Nacional de Genotipado (CeGen-ISCIII; https://xenomica.eu). An additional dataset of Spanish control individuals was provided by the Banco Nacional de ADN Carlos III (BNADN; www.bancoadn.org) and genotyped using the same array at CeGen-ISCIII.
2. RNAseq of blood-derived total RNA, capturing both messenger RNAs (mRNAs) and long non-coding RNAs (lncRNAs), was conducted using the NovaSeq X Plus platform (Illumina). Each sample was sequenced to a depth of approximately 100 million paired-end reads, generating approximately 15 GB of raw data per individual. This high-resolution transcriptomic profiling enables quantification of gene expression, isoform diversity, and the detection of low-abundance transcripts, supporting in-depth analysis of peripheral blood transcriptomes.
3. Epigenomic profiling was performed using the Infinium MethylationEPIC v2.0 BeadChip (Illumina), which interrogates more than 935,000 CpG sites per individual, offering genome-wide coverage of promoter regions, enhancers, and other regulatory elements.

### Computational Infrastructure and Data Processing Environment

The analysis of genomic, transcriptomic, and epigenomic data from the *MadManic* cohort is supported by computational infrastructure that ensures secure data handling, efficient processing, and compatibility with collaborative large-scale projects.

Data are processed using several high-performance computing (HPC):

1. The Scientific Computational Centre (Centro de Computación Científica, CCC) at the Universidad Autónoma de Madrid provides a general-purpose HPC environment used for genome-wide genotyping analysis and statistical genetics.
2. The Drago cluster at CSIC is primarily used for the processing of RNAseq data, and storage of large-scale datasets.
3. The Snellius supercomputer, hosted in the Netherlands, is used in collaboration with the Psychiatric Genomics Consortium (PGC). This cluster supports meta-analytic pipelines and large-scale genomic analysis and serves as the platform where PGC members process their data and share summary statistics for collaborative projects. Snellius supports Ricopili v. 2025_Jan_30.003^38^, which is a computational suite that allows standardized common variant management at all steps: i) pre-imputation QC; ii) principal component analysis; iii) genotype imputation; iv) post-imputation analyses.

## RESULTS

### Biospecimen collection and high throughput arrays

The collection includes over 4,000 participants with more than 11,000 biospecimens, including DNA, RNA, plasma and serum of individuals diagnosed with BD, SCZ, SA, MDD, BPD, SUD, and matched healthy controls (**Table 2**). The collection has been used for a wide range of high-throughput arrays, including genomics, transcriptomics, and epigenetics, enabling integrative multi-omic approaches into disease mechanisms, treatment response, and suicide risk.

Currently, a first SNP genotyping has been completed with the Axiom array, which included 5,785 individuals, being 3,526 from BNADN and 2,259 from the *MadManic* cohort. The Axiom Analysis Suite Software (AxAS, Thermo Fisher Scientific) was used for a first QC approach. After this first filter, 5,767 individuals and 739,777 markers were used for subsequent QC filters at the CCC server. Finally, the pre-imputation module was performed using the Ricopili protocol at the Snellius server. A total of 5,374 samples and 586,001 SNPs underwent genomic imputation with the Haplotype Reference Consortium reference panel^39^, including 21,265 individuals of European ancestry. The imputed genotype dosage is computed into best guess genotypes, which included 7,372,016 SNPs. All QC steps and computational settings are described in **Fig. 3**. A multidimensional scaling analysis (MDS) was performed on 5,548 individuals using the 1000 Genomes Project reference panel,^40^ to visualize superpopulation structure across the *MadManic* cohort (**Fig. 4**). Individuals with East-Asian, African and American ancestry were filtered out for subsequent genomic analyses with European only subjects.

**Figure 3:**
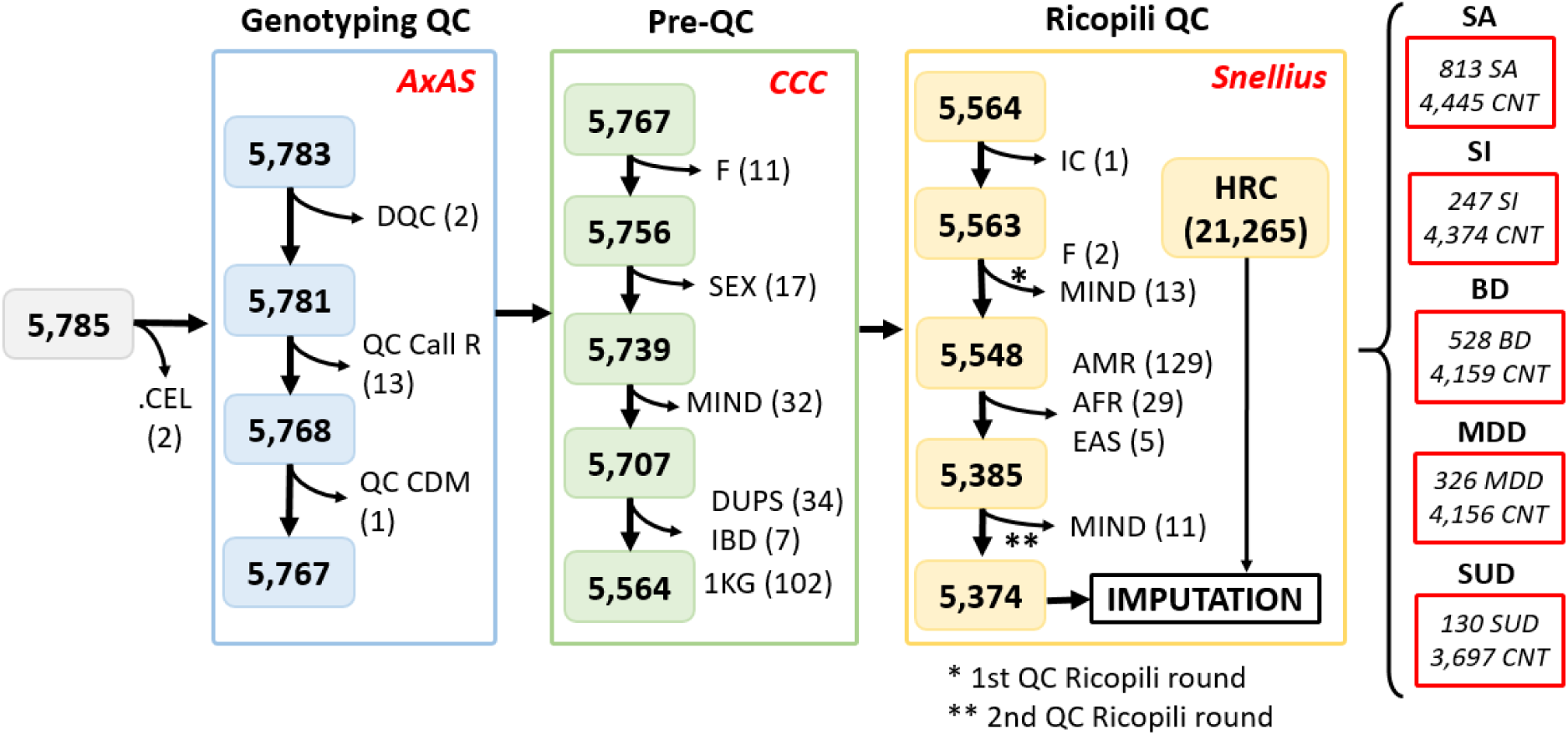
Genotyping, QC workflow and genetic imputation of the *MadManic* cohort. The QC consists of three steps, performed through different platforms (AxAS, CCC and Snellius server). 5,785 individuals were sent for SNP-array genotyping. Subjects in a first QC filtering were discarded for the following reasons: two failed to be genotyped, 16 samples were removed for quality control parameters in AxAS (DQC, QC call R and QC CDM). The second QC step at CCC server (Pre-QC), removed 203 samples for heterozygosity coefficient (F), sex discrepancies (SEX), missing genotype rate (MIND), duplicated individuals (DUPS) and identity by descent analysis (IBD) that identified related individuals. Finally, the QC filters in Snellius via Ricopili pre-imputation module, removed 190 samples, which left only Europeans (5,374 subjects) that were then imputed using the Haplotype Reference Consortium reference panel. Finally, different case-control subsets were generated using different diagnostic criteria: SA (suicide attempt), SI (suicide ideation), BD (bipolar disorder), MDD (major depressive disorder), SUD (substance use disorder). *Abbreviations*: DQC, Dish QC probes; QC call R, call rate of QC probes; QC CDM, Cluster distance mean of QC probes; 1KG, samples belonging to 1000 Genomes project; IC, subject removed for informed consent; AMR, American; AFR, African, EAS, East-Asian; EUR, European; HRC, Haplotype reference consortium.

**Figure 4.**
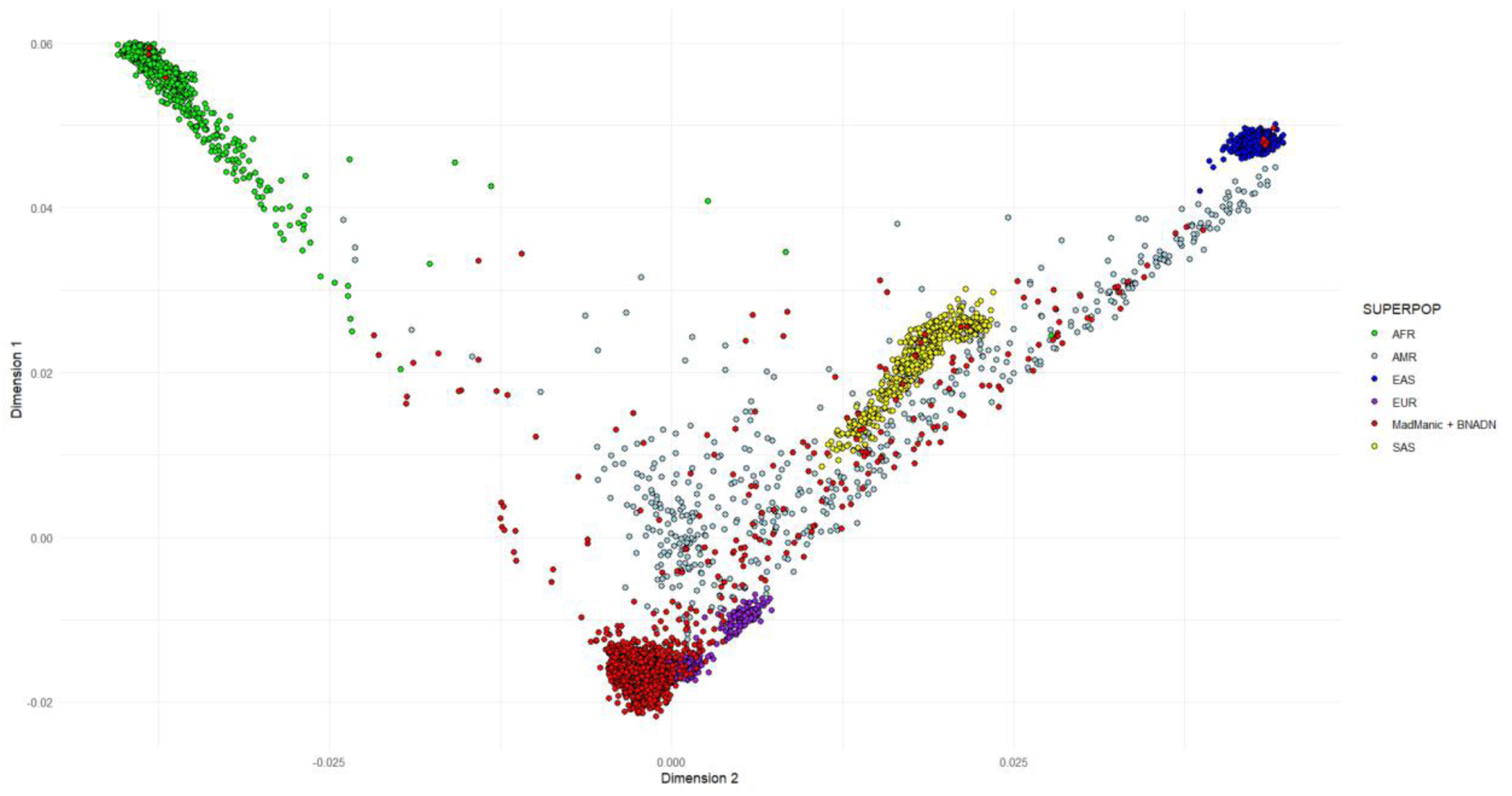
This multidimensional scaling (MDS) plot shows genetic data from our cohorts as red dots (N=5,548 individuals from the *MadManic* group and controls from the BNADN), and 1000 Genome populations. Only the first two MDS components are shown to illustrate genetic clustering of individuals from global populations. *Abbreviations*: AMR, American; AFR, African; SAS, South Asian; EAS, East-Asian; EUR, European.

RNAseq was performed in 725 individuals (444 BD cases, 281 controls) through six different sequencing waves (PILOT=7; W1 = 176; W2= 188; W3=236; WR = 97; WF = 21). Quality sequencing metrics are shown in **Fig. 5**: i) The read length distribution ranged from 141 to 150 base pairs (bp) (**Fig. 5A**); ii) The mean Phred quality score was high (> 39) (**Fig. 5B**); iii)

**Figure 5:**
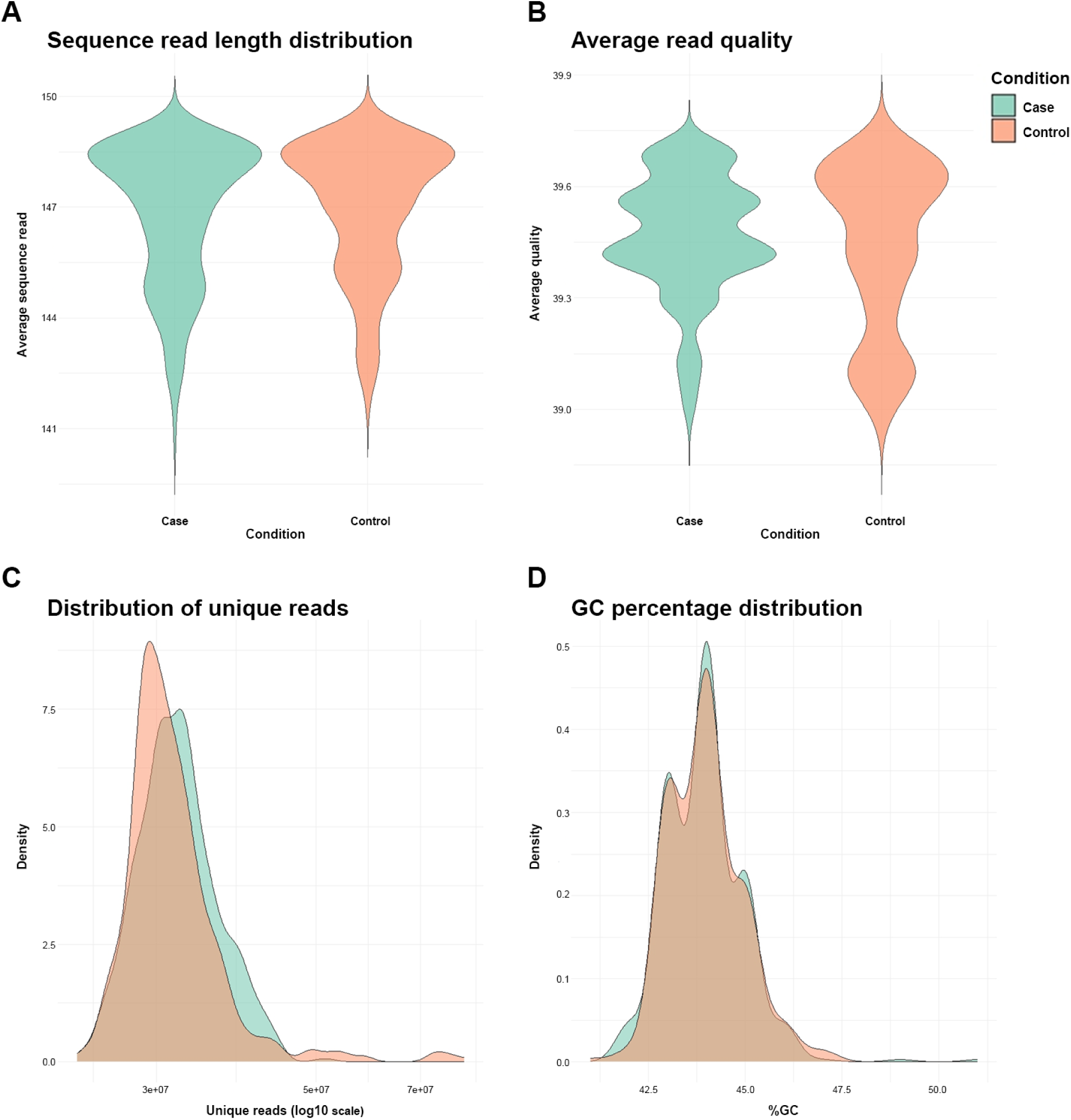
RNAseq quality metrics from 725 individuals of the *MadManic* cohort. **A)** Average sequence read length distribution between groups. Each condition is represented on the x-axis and the distribution of the average read length on the y-axis. **B)** Average read quality (mean Phred scores) distribution per condition group. Each condition is represented on the x-axis and the distribution of the average read quality on the y-axis. **C)** Distribution of unique reads between groups. The unique reads on log10 scale is represented on the x-axis and the distribution of the density on the y-axis. **D)** GC percentage distribution across groups. The GC percentage is represented on the x-axis and the distribution of the density on the y-axis. Cases are shown in green and controls in orange.

Quantification of unique reads per sample were at least 50M reads per sample (**Fig. 5C**); iv) Percentage of guanine-cytosine content (%GC) fell between 42-47% (**Fig. 5D**), in concordance for gene-rich regions in the human genome^41^. Taken together, the RNAseq data obtained is suitable for complex analyses that require a strict QC.

The MethylationEPIC v2.0 array was performed on 239 individuals (130 BD cases and 109 controls). QC was performed following the CPACOR (Control Probe Adjustment and reduction of global CORrelation)^42^ pipeline. Array efficiency and quality parameters are shown in **Fig.6**: i) Bisulfite conversion efficiency (**Fig. 6A**); ii) Sample-level intensities based on control probes (**Fig. 6B**) iii) Raw β-values distribution (**Fig. 6C**). All samples passed basic QC filters, as the mean bisulfite conversion efficiency was 95.58%, and all samples presented a log₂ intensity >10.5, with a median of 11.45. The raw β-values followed the expected distribution, and 825,806 probes were retained for downstream analyses after QC.

**Figure 6:**
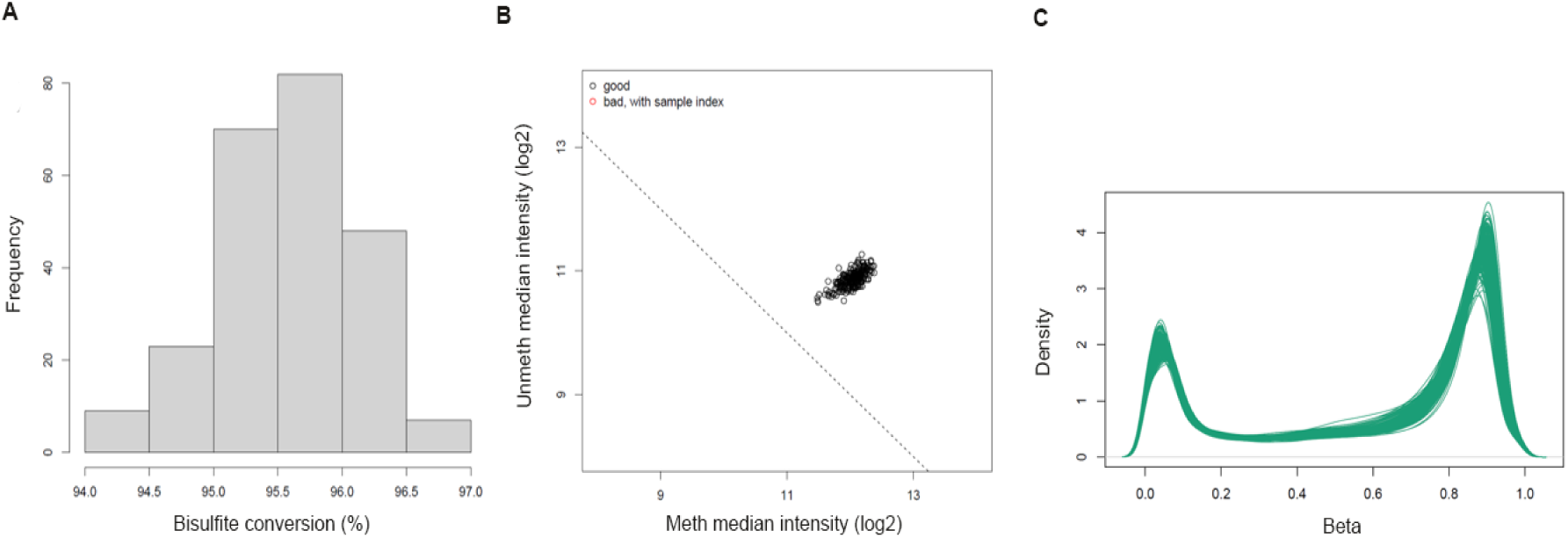
Quality control of for the EPICv2 DNA methylation data from 239 samples of the *MadManic* cohort. **A)** Bisulfite conversion rate (%), showing that all samples overcome the 90% minimum threshold. **B)** Log₂ median intensity of methylated versus unmethylated probes, showing that all samples cluster together overcoming the quality threshold set at 10.5. **C)** Distribution of raw β-values across samples showing expected distribution across all samples.

### Genomic analyses for disease risk and pharmacogenetic pipelines

The imputed genomic data are used to perform GWAS through the Ricopili post-imputation module, and the summary statistics files are shared with the corresponding PGC working groups to contribute with ongoing meta-analysis. Computational pipelines for candidate variants and genes identification include the following approaches^43^: i) Gene-based analysis or gene-set analysis; ii) Gene prioritization framework integrating polygenic risk and multiple biological features; iii) Fine-mapping of GWAS loci to identify putative causal variants.

PRS are derived using PRS-CSx v.1.1.0^44^, which implements a Bayesian continuous shrinkage framework to model the effects of all SNPs jointly. Genetic data and PRS will be analysed with clinical data and phenotypic scales available in the cohort, in order to identify genetic determinants of psychiatric traits or symptoms.

The pharmacogenetics will be explored in two main areas: i) The pharmacogenes of the cytochrome P450 family (*CYP1A2*, *CYP2B6*, *CYP2C9*, *CYP2C19*, *CYP2D6*, *CYP3A4*, and *CYP3A5*); ii) The HLA diplotypes.

The genes of the CYP450 family are responsible for the metabolism of approximately 70–80% of clinically used drugs. Common and rare variants from the Axiom genomic array are analysed through pyPGX^45^, AlleleTyper (Thermo Fisher), and Genostar^46^, which primarily rely on the ClinPGx database (https://www.clinpgx.org/) to infer metabolizer phenotypes.

HLA diplotypes will be inferred from genomic data using the Axiom HLA Analysis software (Applied Biosystems). Haplotypes from the HLA genes are associated with drug hypersensitivity reactions and other immune-mediated adverse effects observed with carbamazepine, a mood stabilizer used to treat bipolar disorder^47^.

The pharmacogenetic profile obtained from these analyses will be shared with psychiatrists to facilitate translation into clinical practice.

### Transcriptomic approaches for gene expression, co-expression networks and splicing events

RNAseq data are used for differential gene expression (DGE) analyses based on the affection status or pharmacological treatment at the time of sample extraction. Weighted Gene Co-expression Network Analyses (WGCNA) are applied to identify modules or hubs of highly co-expressed genes that can be subsequently correlated with primary psychiatric diagnosis, medication response, and quantitative clinical scores to enable the identification of biologically related gene sets associated with the trait.

The CIBERSORTx pipeline is used to infer the expression profile of 22 immune cell types^48^. These results will be used to assess potential relations between psychiatric conditions and medication with cell-type proportions, which is known in the case of lithium to increase white blood cell count, especially neutrophils^49^.

Transcriptome-wide alternative splicing analyses (TWASA) are performed using MAJIQ v.3, which quantifies both basic and complex alternative splicing events by modelling junction-level read support to define local splicing variations (LSVs) and estimate changes in splice-junction usage across samples^50^. Additionally, aberrant splicing detection is implemented via FRASER 2.0 to identify individual-level transcript isoforms^51^.

The imputation of gene expression in brain tissues is performed through BrainGENIE that uses peripheral blood transcriptomic data to predict brain tissue-specific gene expression^52^. This methodology can be further applied to DGE and WGCNA analyses, facilitating the investigation of neural alterations associated with psychiatric conditions.

### Epigenome-wide association studies, polymethylation score and epigenomic clocks

The beta values obtained after QC in the EPICv2 array are transformed into M-values for downstream Epigenome-Wide Association Studies (EWAS). Adjusted M-values are then regressed on case–control status to identify genome-wide differentially methylated positions (DMPs) associated with the trait.

Furthermore, the EWAS summary statistics can be used to identify Differentially Methylated Regions (DMRs)^53^. DMRs are defined as clusters of significant or suggestive CpGs located within a window. While individual CpGs may show only modest effects, grouping them into DMRs increases power to detect coordinated methylation changes, which may have an effect in gene expression and can be subsequently validated with RNAseq data.

Residualized autosomal beta values are used to identify Co-Methylated Regions (CMRs) with the CoMeBack R package^54^. CMRs are defined as CpG groups located within a window of 500bp that show pairwise correlations above 0.3. The Poly-Methylation Score (PMS), similarly to PRS, can also be derived from the EWAS summary statistics for certain traits. Methylation scores are then computed across multiple P-value thresholds, and the PMS is defined by their first principal component^55^.

Epigenetic clocks based on the beta values can be used to estimate epigenetic age based on methylation levels at specific CpGs. The *methylCIPHER* R package is used to implement several clocks, including *PhenoAge*, which reflects morbidity-related aging, and *GrimAge*, which reflects mortality risk^56–58^. Epigenetic age acceleration is defined as the residual from modelling epigenetic age as a function of chronological age.

### Integrative multi-omics approaches to uncover functional genetic mechanisms

We aim to integrate multiple layers of genomic and transcriptomic data to enable novel, functionally informed analyses for psychiatric disorders. Transcriptome-wide association studies (TWAS) combine common variants (SNPs) with gene expression data to identify genes whose expression is associated with disease risk, thereby moving from SNP-level signals to more interpretable gene-level mechanisms^59,60^. Complementarily, allele-specific expression (ASE) analysis leverages both genotype and RNAseq data to quantify allelic imbalance at heterozygous sites, providing direct insight into cis-regulatory effects on gene expression^61^. Together, these approaches that we will develop for our cohort, will enhance the identification of candidate causal genes and regulatory mechanisms, offering a more comprehensive understanding of the molecular pathways underlying psychiatric phenotypes.

## DISCUSSION

Current international initiatives in psychiatry are focused on unravelling the complex architecture between genetic markers and clinical phenotypes. To advance this field, it is imperative to establish large-scale, deeply phenotyped cohorts that bridge comprehensive clinical profiles with multi-omic data. Such integration is crucial for improving risk stratification, developing preventive frameworks, and enabling precision interventions.

Here, we present the *MadManic* cohort as a comprehensive and integrative resource for investigating SMDs, including BD, SCZ, and MDD, with a particular focus on suicidality and treatment-related outcomes. The design of this cohort responds to a critical need in psychiatric research: the integration of large-scale genomic data with deep clinical phenotyping, digital monitoring, and multi-omic approaches to better capture the complexity and heterogeneity of these disorders. These approaches are desirable given the estimations of large number of risk alleles implicated in SMDs, and their interaction with the environment^17^.

One of the main strengths of the *MadManic* cohort lies in its transdiagnostic framework, enabling the systematic comparison of genetic risk profiles across BD, SCZ, and MDD^62^. Given the substantial genetic overlap reported among psychiatric disorders, this approach facilitates the identification of both shared and disorder-specific biological mechanisms^63^. By leveraging genome-wide data and PRS, this cohort will provide a platform to investigate how genetic liability contributes not only to disease risk but also to clinical variability, including symptom severity, disease trajectory, and suicidal behaviour^16^.

Importantly, the integration of genetic data with standardized clinical scales, as those included in our digital clinical protocol, allows for a more refined characterization of phenotype and risk alleles relationships^64^. In addition, the inclusion of neurobiological proxies such as laterality, assessed through the Edinburgh Handedness Inventory (EHI), offers novel opportunities to explore neurodevelopmental dimensions of psychiatric disorders^65^. The higher prevalence of non-right handedness reported in SCZ compared to controls may reflect underlying neurobiological differences that can now be interrogated in conjunction with genomic data within this cohort^66^.

Another key aspect of the *MadManic* cohort is the incorporation of digital phenotyping through EMA, enabling the collection of longitudinal, real-time behavioural and clinical data^13,14^. This represents a major advancement over traditional cross-sectional assessments and opens new avenues for dynamic risk prediction, particularly in relation to suicidal behaviour. The combination of longitudinal digital data with genomic information is expected to significantly improve predictive modelling through machine learning approaches, moving the field closer to clinically actionable tools^67^.

Furthermore, this cohort is uniquely positioned to contribute to international efforts in psychiatric genetics. The contribution of *MadManic* cohort to the PGC will substantially increase the representation of Southern European populations in global meta-analyses. This is particularly relevant given the current underrepresentation of non-Northern European ancestries in large-scale genomic studies, which may limit the generalizability of findings and the equitable implementation of precision psychiatry^68^.

While individual genomic, transcriptomic, or epigenomic datasets provide critical insights, they often capture only a fraction of the complex risk architecture underlying psychiatric conditions. Recent bioinformatic advances now allow for the integrative combination of these different biological layers, offering an unprecedented opportunity to map specific compromised pathways unique to each psychiatric disorder.

Building upon our established datasets of genomic, transcriptomic, and epigenomic profiling, our future research will aim to integrate also untargeted metabolomic and proteomic arrays. The inclusion of these additional layers will contribute to bridge existing gaps in the molecular landscape of psychiatry. By transitioning from isolated genetic data to a comprehensive multi- omic framework, we expect to uncover critical biological mechanisms that remain unexplored when analysed through a single lens.

Despite these strengths, several limitations should be acknowledged. First, as recruitment is ongoing, there is currently an imbalance in sample sizes across diagnostic groups, particularly for SCZ, which may limit the power of certain comparative analyses. Efforts are underway to expand the recruitment of SCZ patients to address this limitation. Second, while the use of clinical scales and digital tools enhances phenotypic resolution, some measures rely on self-report or routine clinical assessments, which may introduce variability or bias.

Additionally, although the project aims to integrate multi-omic data, including transcriptomics and other functional genomic layers, these datasets are still being generated and expanded. The full potential of integrative analyses, such as TWAS and ASE, will be performed in subsequent phases of the project. These approaches are expected to provide critical functional insights into GWAS findings and help unravel the complex relation between genetic associations and biological mechanisms.

In conclusion, the *MadManic* cohort will allow significant step forward in psychiatric research by combining large-scale genomic data, deep clinical phenotyping, digital monitoring, and emerging multi-omic approaches within a single, well-characterized cohort. This integrative framework has the potential to advance our understanding of the biological basis of SMDs, improve risk stratification, and ultimately contribute to the development of personalized interventions in psychiatry.

## DATA AVAILABILITY

Individual-level data are not publicly available for data sharing. Sharing of summary data will be made available upon request.

## ACKNOWLEDGMENTS

We would like to express our deepest gratitude to all patients and volunteers who participated in this study. We also thank Ivan de la Calle González, CEO of “*Cobaro Soluciones*” for the technical support and implementation of the digital protocol, Dominic Petru Demeterca Ungureanu for advanced support in automation of analytical pipelines, and Mónica Campos García for her involvement in controls recruitment. We also thank the Banco Nacional de ADN Carlos III (BNADN) (www.bancoadn.org) for providing part of the genotyping data of our control sample, and the “Centro Nacional de Genotipado-Fundación Pública Galega de Medicina Xenómica” for the genotyping service.

## AUTHOR CONTRIBUTION

CT and EBG conceptualized and designed the study cohort. Statistical and computational analyses were performed by IGO, MMJ, ICL. Recruitment of patients was carried out by RABS, SSA, SBL, LMI, RAG, EDS, AJO, ALF, IAC, AMGB, while recruitment of controls was carried out by IGO, RSC, MJSA, and CT. Phenotyping and clinical data was assessed by EMP, LJM, ALF, LAG. Extraction of blood samples was done by RABS, ICG, JAV, EAB and RJP, with medical oversight by MBB, SBL, LMI, RAG, and EDS. Recruitment scheduling and organization was performed by CLH, DRM, and LAG. Lab processing was performed by RSC, MMJ, SFR, CVL, NBG, PH. The digital clinical protocol was designed by CT, SSA, JMF, LAG, and EBG. Funding was obtained by CT and EBG. IGO and CT contributed substantially to drafting the manuscript. All authors contributed to review the manuscript and approved the final version.

