## Supplementary Table S1 for "The Madrid Manic Group (MadManic) Cohort: Multi-Omics and Digital Phenotyping For the Studies of Severe Mental Disorders and Suicidality"

**Table S1**: Pharmaceutical drugs registered for BD patients in the MadManic cohort.

| Category | Generic name | Trade name |
|---|---|---|
| **Antidepressants** | | |
| SSRIs | Fluoxetin | Prozac, Adofen |
| | Sertralin | Aremis, Besitran, Altisben |
| | Citalopram | Prisdal, Seropram |
| | Escitalopram | Cipralex, Esertia |
| | Paroxetine | Seroxat, Motivan, Frosinor, Daparox |
| | Fluvoxamine | Dumirox |
| SNRIs | Venlafaxine | Vandral, Dobupal |
| | Desvenlafaxine | Pristic |
| | Duloxetine | Cymbalta, Dulotex, Xeristar |
| TCAs | Amitriptyline | Nobritol, Triptizol |
| | Clomipramine | Anafranil |
| | Imipramine | Tofranil |
| | Nortriptyline | Paxtibi |
| | Trimipramine | Surmontil |
| | Doxepin | Sinequan |
| | Maprotiline | Ludiomil |
| Other | Mirtazapine | Rexer |
| | Bupropion | Elontril |
| | Reboxetine | Irenor, Norebox |
| | Atomoxetine | Strattera |
| | Vortioxetine | Brintellix |
| **Antipsychotic** | | |
| | Paliperidone | Invenga |
| | Risperidone | Risperdal |
| | Aripiprazole | Abilify |
| | Quetiapine | Seroquel, Psicotric |
| | Asenapine | Sycrest |
| | Olanzapine | Zyprexa |
| | Haloperidol | Haloperidol |
| | Amisulpride | Solian |
| | Lurasidone | Latuda |
| | Ziprasidone | Zeldox |
| | Cariprazine | Reagila |
| **MAOI** | | |
| | Phenelzine | Nardil |
| | Selegiline | Emsam |
| | Isocarboxazid | Marplan |
| | Tranylcypromine | Parnate |
| **Mood stabilizers** | | |
| | Lithium | Plenur |
| | Valproic acid | Depakine |
| | Lamotrigine | Lamictal |
| | Carbamazepine | Tegretol |
| | Oxcarbazepine | Trileptal |
| | Gabapentin | Neurontin |
| | Pregabalin | Lyrica |

| Anxiolytics/sedatives | | |
|---|---|---|
| | Benzodiazepines | Orfidal, Lexatin, Rivotril Valium, Tranxilium, Noctamid |

SSRIs *Selective Serotonin Reuptake Inhibitors*; SNRIs *Serotonin-Norepinephrine Reuptake Inhibitors*; TCAs *Tricyclic Antidepressants*; MAOI *Monoamine Oxidase Inhibitor*
